# Factors associated with human papillomavirus, hepatitis A, hepatitis B and mpox vaccination uptake among gay, bisexual and other men who have sex with men in the UK – findings from the large community-based RiiSH-Mpox survey

**DOI:** 10.1101/2024.03.08.24303981

**Authors:** George Baldry, Dawn Phillips, Ruth Wilkie, Marta Checchi, Kate Folkard, Ruth Simmons, John Saunders, Sema Mandal, Catherine H Mercer, Hamish Mohammed, Dana Ogaz

## Abstract

**Background:** Gay, bisexual, and other men who have sex with men (GBMSM) face a disproportionate burden of sexually transmitted infections and are eligible for targeted vaccinations for hepatitis A (HAV), hepatitis B (HBV), human papilloma virus (HPV) and mpox. This study examines the sociodemographic characteristics, sexual behaviours, and sexual healthcare service (SHS) use associated with vaccination uptake.

**Methods:** We undertook analyses of RiiSH-Mpox - an online, community-based survey with GBMSM recruited via social media and dating apps. We calculated vaccination uptake (≥1 dose) among eligible GBMSM. Bivariate and multivariable logistic regression was performed to identify factors independently associated with vaccination uptake among eligible participants.

**Results:** Reported uptake in eligible GBMSM was around two-thirds for each of the vaccinations considered: mpox 69% (95% confidence interval (CI): 66%-72%), HAV 68% (CI:65%-70%), HBV 72% (CI:69%-74%) and HPV 65% (CI:61%-68%). Vaccination course completion (receiving all recommended doses) ranged from 75% (HBV) to 89% (HAV) among eligible GBMSM. Individuals who represented missed opportunities for vaccination ranged from 22-30% of eligible SHS attendees. Younger participants, individuals identifying as bisexual, reporting lower educational qualifications, or being unemployed reported lower uptake across GBMSM-selective vaccinations. Individuals who reported greater levels of sexual behaviour and recent SHS use were more likely to report vaccinations.

**Conclusion:** Eligible participants reported high uptake of vaccinations; however, uptake was lower amongst young GBMSM and self-identifying bisexual men. Awareness of groups with lower vaccination uptake will help inform practice, delivery strategies and health promotion, to improve the reach and impact of vaccinations amongst GBMSM.

**Contributions:** DP, RW, KF, CHM, JS, HM reviewed and updated the survey instrument. Survey implementation, data collection and data management were carried out by RW and DO. GB, CM, DO, HM curated secondary analysis plan with review and contributions from KF, MC, JS, RS, SM. GB conducted analysis and completed the first manuscript draft. All authors contributed to successive drafts and reviewed and approved the final manuscript.

**Ethics statement:** Ethical approval of this study was provided by the UKHSA Research and Ethics Governance Group (REGG; ref: R&D 524). Online informed consent was received from all participants and all methods were performed in accordance with guidelines and regulations set by the UKHSA REGG.

**Competing interests:** Authors have no competing interests to declare.

**Data availability statement:** The data that support the findings of this study are available upon reasonable request from the UK Health Security Agency (UKHSA). Requests can be directed to Dr Hamish Mohammed.

**Funding:** This study did not receive any funding and was conducted as part of the UKHSA public health response to mpox.

## Introduction

Gay, bisexual, and other men who have sex with men (GBMSM) continue to be disproportionately affected by sexually transmitted infections (STIs) and viral hepatitides (VHs) both globally^1^ and in the UK^2^. To reduce individual and public health impacts of these infections in the UK, several vaccines are recommended for GBMSM^3–6,^ this includes HPV (human papilloma virus), Hepatitis A (HAV) and Hepatitis B (HBV), which can be spread through sexual contact amongst other means^7^.

UK immunisation policy and clinical advice is outlined in The Green Book: Immunisation against Infectious Disease, a compilation of immunisation evidence, procedures, and guidance for vaccine preventable diseases^8^. The UK currently recommends opportunistic immunisation against HAV and HBV to all GBMSM, particularly those who change sexual partners frequently^3,7^. HPV vaccination in the UK was initially introduced for girls aged 12-13 to prevent cervical cancer^9,^ but was recommended for GBMSM up to and including age 45 from 2016 and for adolescent boys from 2019^5^. Most recently, GBMSM-selective vaccination programmes have been brought into focus through the rapid introduction of vaccination for mpox in response to the 2022 mpox outbreak which particularly affected this group^10^.

There is limited literature examining STI/VH vaccination uptake and acceptability among GBMSM. While a pilot study among UK GBMSM found that 89% would willingly receive the HPV vaccine upon recommendation^11^, reported uptake of the HPV vaccine is only 34% among GBMSM attending sexual health services (SHS) in England (uptake has also likely decreased due to the impact of COVID-19 on SHS in-person service delivery)^12^. Data on HAV/HBV vaccination uptake among GBMSM in England is also very limited.

To understand STI/VH vaccination uptake inequalities, we investigated the factors associated with uptake of recommended STI/VH vaccines in the UK among a community-sample of GBMSM.

## Methods

### Study design and Data collection

This study used data collected by the 2022 round of the ‘Reducing inequalities in Sexual Health during the mpox outbreak’ (RiiSH-mpox) survey, part of a series of cross-sectional community-based surveys first run in 2017^13^. The RiiSH surveys focus on health and wellbeing, sexual risk behaviours, and use of SHS amongst GBMSM, and have been used to improve understanding of the factors underlying STI trends among GBMSM in the UK^14,15^. RiiSH-Mpox included questions on these core topics as well as mpox vaccine uptake during the 2022 mpox outbreak. Details of the RiiSH-Mpox methodology have been previously reported^15^.

The RiiSH-Mpox survey was deployed from 24 November-19 December 2022 and included participants that were aged ≥16 years, a UK resident, individuals self-identifying as men (cisgender/transgender), transgender women, or gender-diverse individuals assigned male at birth (gender was overlooked in this analysis, 99% self-identified as a cis-male), and individuals who self-reported having had sex with a man in the last year. Participants were recruited through advertisements on social media (Facebook, Instagram, Twitter [now X]) and through the geospatial networking application (‘dating app’) Grindr. Online consent was provided by all participants. Lookback periods for healthcare-related factors and sexual risk behaviours varied from reporting since August 2022 (i.e. last 3-4 months), to ever.

Due to relatively low numbers of participants who did not self-report being white or gay/homosexual these variables were dichotomised as white or any other ethnic group, and either gay/homosexual or bisexual, respectively.

### Data analysis

#### Vaccination uptake

To measure uptake and completion levels of vaccination, percentages and 95% confidence intervals (CI) (Clopper-Pearson) were calculated. Uptake was defined as receiving at least one dose of a vaccine and completion as receiving all recommended doses as outlined by the UK vaccination guidance at the time of the survey (2 doses of HAV, HPV or mpox vaccine and 3 HBV vaccine doses).

Uptake amongst all participants and eligible GBMSM for each vaccine was calculated. Reflecting UK vaccination guidance ^3^, we considered all GBMSM as eligible for HAV and HBV vaccination. HPV vaccination eligibility was considered in GBMSM reporting SHS attendance who were up to and including age 45, given opportunistic vaccination recommendations across SHS. In line with mpox vaccination guidance^6,10^, mpox vaccination eligibility was considered in those reporting: ≥10 physical male sex partners, meeting partners in public sex environments (Sex on premise venues (SOP), cruising grounds or sex parties), having a positive STI test, report of HIV-PrEP or use of chemsex-associated recreational drugs (crystal methamphetamine, mephedrone or gamma-hydroxybutyrate/gamma-butyrolactone). SHS attendance was not considered as part of eligibility for HAV, HBV and mpox vaccination given vaccination availability outside of SHS (e.g. mpox vaccination outreach, general practice and travel clinic availability for VH vaccination).

To measure the association between uptake of different vaccine types, logistic regression was used to generate the unadjusted (uOR) and adjusted odds ratio (aOR) of receiving at least one dose of a respective vaccine versus also receiving at least one dose of another vaccine (vaccines were compared sequentially, in a pairwise manner). For adjusted analyses, age-group and sexual orientation were considered a priori, and additional factors were added in a forwards stepwise manner based on significant bivariate association and evidence from previous literature^11,16–19^.

#### Missed vaccination opportunities in those attending sexual health services

GBMSM who reported ever having attended an SHS and who met respective proxy vaccination eligibility were considered missed opportunities for vaccination if no vaccination history was reported.

#### Factors associated with vaccination uptake

Among all participants irrespective of previous SHS attendance (except those over age 45/ those who had not previously attended SHS for HPV vaccination), Pearson’s chi-squared test and bivariate logistic regression was used to examine association between sociodemographic, sexual behaviour and health-related factors with vaccination uptake for the HAV, HBV and HPV vaccines, respectively. Analysis of factors associated with vaccination for mpox uptake has been previously reported^15^ so was not examined in this analysis. Multivariable adjustment for significant sociodemographic, sexual behaviour and health-related factors, as per methods described above, was applied.

All analysis was completed using Stata V17.0 (College Station, TX, USA) and all p values below 5% were deemed statistically significant.

## Results

Altogether, 1435 individuals engaged with RiiSH-Mpox survey, of whom 1333 met the inclusion criteria. Of these, half were aged over 45 years, 37% were 30-44 and the remaining 13% were 16-29. Almost all self-identified as gay/homosexual (89%) and were of white ethnicity (92%). Most lived in England (86%), 81% were employed, 37% reported at least degree level education, with 48% responding that they were financially comfortable. See Appendix A for sample characteristics.

### Vaccination Uptake

Among all participants, 97% self-reported receiving at least one dose of any STI/VH vaccination. HAV vaccination uptake was 68% (CI:65%-70%, 903/1333) and HBV vaccination was 72% (CI:69%-74, 954/1333). Mpox vaccination uptake among eligible participants was 69% (CI:66%-72%, 601/875), or 52% (CI:49%-55%, 692/1,333) among all GBMSM. In those under age 45 who had ever attended a SHS, HPV vaccination was 73% (CI:69%-76%, 417/574), or 42% (CI:39%-45%, 562/1333) among all participants.

Receiving one vaccine type was uniformly associated with receiving at least one other (Appendix B). The highest association was found between the HAV and HBV vaccines, individuals who had received one were 12 times more likely to have received at least one dose of the other (aOR:11.98, CI: 9.01-15.92).

### Vaccine Completion

HAV vaccination had the highest completion level amongst participants with 89% (CI:87%-91%, 807/903) of those that had received at least one dose completing the course. Among individuals that initiated HBV vaccination, 75% completed the course (CI:72%-77%, 712/954). Of those that had initiated HPV vaccination 84% (CI:80%-88%, 350/ 417) had completed the course and 42% (CI:38%-45%, 288/692) of those that had initiated vaccination for mpox completed the course.

### Missed vaccination opportunities in those attending sexual health services

Missed vaccination opportunities among GBMSM who ever reported SHS attendance history were considered 27% (CI:24-29%) for HAV vaccination, 22% (CI:20-25%) for HBV vaccination, 27% (CI: 26-33%) for HPV vaccination, and 30% (CI: 26-33%) for mpox vaccination.

### Factors associated with vaccination uptake

#### Hepatitis A Vaccination uptake

There was a bivariate association between HAV uptake and sexual orientation, educational level, and employment status, see Table 2. There were significantly lower odds of HAV vaccination amongst those self-reporting as bisexual (aOR:0.56, (CI:0.4-0.8) vs gay/homosexual), having an education that was below degree-level (aOR:0.7, (CI:0.55-0.9)) and being unemployed (aOR:0.69, (CI:0.52-0.92)). There was no evidence of an independent association existing between HAV vaccination uptake and age, however uptake was lower in both the 16-29 age group (aOR:0.54, (CI:0.37-0.78)) and the over 45 age group (aOR:0.74, (CI:0.57-0.97)).

**Table 2.** Sociodemographic characteristics and Health-related and sexual behaviour factors associated with self-reported HAV vaccination uptake among RiiSH-Mpox participants.

|  | n total | % of RiSH-Mpox participants | n of HAV vaccinated participants * | % of group vaccinated | % of HAV vaccinated people | uOR ** | 95% confidence interval | P | aOR § | 95% confidence interval | P |
| --- | --- | --- | --- | --- | --- | --- | --- | --- | --- | --- | --- |
| **Sociodemographic Characteristics** |  |  |  |  |  |  |  |  |  |  |  |
| **Age-group** |  |  |  |  |  |  |  |  |  |  |  |
| 16-29 | 176 | 13 | 101 | 57 | 11 | 0.48 | 0.34 - 0.69 |  | 0.54 | 0.37 - 0.78 | 0.570 |
| 30-44 | 486 | 36 | 358 | 74 | 40 | 1.00 |  |  | 1.00 |  |  |
| 45+ | 671 | 50 | 444 | 66 | 49 | 0.70 | 0.54 - 0.90 | 0.570 | 0.74 | 0.57 - 0.97 |  |
| **Sexual orientation** |  |  |  |  |  |  |  |  |  |  |  |
| Gay/ homosexual | 1187 | 89 | 823 | 69 | 91 | 1.00 |  |  | 1.00 |  |  |
| Bisexual † | 146 | 11 | 80 | 55 | 9 | 0.54 | 0.38 - 0.76 | <0.001 | 0.56 | 0.40 - 0.80 | 0.001 |
| **Ethnicity** |  |  |  |  |  |  |  |  |  |  |  |
| White | 1223 | 92 | 832 | 68 | 92 | 1.00 |  |  | 1.00 |  |  |
| Other | 110 | 8 | 71 | 65 | 8 | 0.86 | 0.57 - 1.29 | 0.454 | 0.90 | 0.59 - 1.37 | 0.610 |
| **Country of Birth** |  |  |  |  |  |  |  |  |  |  |  |
| UK | 1113 | 83 | 756 | 68 | 84 | 1.00 |  |  | 1.00 |  |  |
| Other | 220 | 17 | 147 | 67 | 16 | 0.95 | 0.70 - 1.29 | 0.748 | 0.91 | 0.66 - 1.26 | 0.586 |
| **Country of Residence** |  |  |  |  |  |  |  |  |  |  |  |
| England | 1148 | 86 | 779 | 68 | 86 | 1.00 |  |  | 1.00 |  |  |
| Rest of UK | 185 | 14 | 124 | 67 | 14 | 0.96 | 0.69 - 1.34 | 0.823 | 0.98 | 0.70 - 1.37 | 0.904 |
| **Educational level** |  |  |  |  |  |  |  |  |  |  |  |
| Degree level or higher | 495 | 37 | 594 | 71 | 66 | 1.00 |  |  | 1.00 |  |  |
| Below degree level | 838 | 63 | 309 | 62 | 34 | 0.68 | 0.54 - 0.86 | 0.001 | 0.70 | 0.55 - 0.90 | 0.050 |
| **Employment** |  |  |  |  |  |  |  |  |  |  |  |
| Currently Employed | 1074 | 19 | 749 | 70 | 83 | 1.00 |  |  | 1.00 |  |  |
| Not Employed | 259 | 81 | 154 | 59 | 17 | 0.64 | 0.48 - 0.84 | 0.002 | 0.69 | 0.52 - 0.92 | 0.012 |
| **Household Composition** |  |  |  |  |  |  |  |  |  |  |  |
| Alone | 506 | 38 | 552 | 67 | 61 | 1.00 |  |  | 1.00 |  |  |
| With Others | 827 | 62 | 351 | 69 | 39 | 0.87 | 0.70 - 1.12 | 0.321 | 0.88 | 0.69 - 1.12 | 0.307 |
| **Relationship Status** |  |  |  |  |  |  |  |  |  |  |  |
| Relationship | 591 | 44 | 510 | 86 | 56 | 1.00 |  |  | 1.00 |  |  |
| Single | 742 | 56 | 393 | 53 | 44 | 1.11 | 0.88 - 1.40 | 0.870 | 1.08 | 0.85 - 1.37 | 0.540 |
| **Comfortable Financial Status** |  |  |  |  |  |  |  |  |  |  |  |
| Yes †† | 693 | 52 | 453 | 65 | 50 | 1.00 |  |  | 1.00 |  |  |
| No | 640 | 48 | 450 | 70 | 50 | 0.80 | 0.63 - 1.00 | 0.054 | 0.93 | 0.73 - 1.18 | 0.551 |
| **Health-related factors** |  |  |  |  |  |  |  |  |  |  |  |
| **HIV Status** |  |  |  |  |  |  |  |  |  |  |  |
| Negative/ Unknown | 1131 | 85 | 744 | 66 | 82 | 1.00 |  |  | 1.00 |  |  |
| PLWHIV | 202 | 15 | 159 | 79 | 18 | 1.92 | 1.34 - 2.75 | <0.001 | 2.00 | 1.38 - 2.88 | <0.001 |
| **MPOX-related behaviour modification ¶** |  |  |  |  |  |  |  |  |  |  |  |
| No | 626 | 47 | 412 | 66 | 46 | 1.00 |  |  | 1.00 |  |  |
| Yes | 707 | 53 | 491 | 69 | 54 | 1.18 | 0.94 - 1.49 | 0.157 | 1.11 | 0.88 - 1.42 | 0.355 |
| **STI Positive Test** |  |  |  |  |  |  |  |  |  |  |  |
| No | 1177 | 70 | 778 | 66 | 86 | 1.00 |  |  | 1.00 |  |  |
| Yes | 156 | 30 | 125 | 80 | 14 | 3.38 | 2.63 - 4.35 | <0.001 | 1.95 | 1.29 - 2.96 | 0.002 |
| **Visited SHC in past year** |  |  |  |  |  |  |  |  |  |  |  |
| No | 478 | 36 | 221 | 46 | 24 | 1.00 |  |  | 1.00 |  |  |
| Yes | 855 | 64 | 682 | 80 | 76 | 4.58 | 3.59 - 5.86 | <0.001 | 4.31 | 3.36 - 5.54 | <0.001 |
| **STI test in past year** |  |  |  |  |  |  |  |  |  |  |  |
| No | 405 | 30 | 196 | 48 | 22 | 3.41 |  |  | 1.00 |  |  |
| Yes | 928 | 70 | 707 | 76 | 78 | 1.00 | 2.67 - 4.37 | <0.001 | 3.25 | 2.53 - 4.18 | <0.001 |
| **PrEP Use in Past Year** |  |  |  |  |  |  |  |  |  |  |  |
| No | 734 | 55 | 415 | 57 | 46 | 1.00 |  |  | 1.00 |  |  |
| Yes | 599 | 45 | 488 | 81 | 54 | 3.38 | 2.63 - 4.35 | <0.001 | 3.15 | 2.43 - 4.07 | <0.001 |
| **Sexual behaviours** |  |  |  |  |  |  |  |  |  |  |  |
| **Usage of chemsex-related drugs in the previous year** |  |  |  |  |  |  |  |  |  |  |  |
| No | 1222 | 92 | 815 | 67 | 90 | 1.00 |  |  | 1.00 |  |  |
| Yes | 111 | 8 | 88 | 79 | 10 | 1.91 | 1.19 - 3.07 | 0.007 | 1.87 | 1.16 - 3.02 | 0.010 |
| **Number of Male Physical Sex Partners** |  |  |  |  |  |  |  |  |  |  |  |
| none | 100 | 8 | 64 | 64 | 7 | 1.00 |  |  | 1.00 |  |  |
| 1 | 180 | 14 | 93 | 52 | 10 | 0.60 | 0.36 - 0.99 |  | 0.59 | 0.35 - 0.98 | <0.001 |
| 2 to 4 | 389 | 29 | 256 | 66 | 28 | 1.08 | 0.68 - 1.71 |  | 1.05 | 0.66 - 1.68 |  |
| 5 to 9 | 280 | 21 | 192 | 69 | 21 | 1.23 | 0.76 - 1.98 |  | 1.17 | 0.72 - 1.90 |  |
| 10+ | 384 | 29 | 298 | 78 | 33 | 1.95 | 1.21 - 3.13 | 0.006 | 1.78 | 1.10 - 2.88 |  |
| **reported use of SOP venues in the lookback period** |  |  |  |  |  |  |  |  |  |  |  |
| No | 888 | 67 | 568 | 64 | 63 | 1.00 |  |  | 1.00 |  |  |
| Yes | 445 | 33 | 335 | 75 | 37 | 1.72 | 1.33 - 2.21 | <0.001 | 1.58 | 1.22 - 2.05 | <0.001 |
| **Number of CAS partners in the lookback period (since Dec 2021)** |  |  |  |  |  |  |  |  |  |  |  |
| none | 401 | 30 | 245 | 61 | 27 | 1.00 |  |  | 1.00 |  |  |
| 1 | 263 | 20 | 159 | 60 | 18 | 0.97 | 0.71 - 1.34 |  | 0.94 | 0.68 - 1.29 | <0.001 |
| 2 to 4 | 315 | 24 | 219 | 70 | 24 | 1.45 | 1.06 - 1.99 |  | 1.42 | 1.03 - 1.95 |  |
| 5 to 9 | 146 | 11 | 118 | 81 | 13 | 2.68 | 1.70 - 4.24 |  | 2.42 | 1.52 - 3.85 |  |
| 10+ | 208 | 16 | 162 | 78 | 18 | 2.24 | 1.53 - 3.29 | <0.001 | 2.1 | 1.42 - 3.10 |  |

\* Participants with self-reported uptake of ≥1 HAV vaccine dose(s). † Includes those identifying as bisexual, straight, or another way. †† Top two quartiles ("I am comfortable"/"I am very comfortable" from the question, "How would you best describe your current financial situation". ¶ Behaviour modification includes self-report of any of the following from May 2022: fewer sexual partners, reduced visits to sex on premises venues or PSE (i.e. cruising grounds), and avoiding: all sex, condomless anal sex, skin-to-skin contact, or visiting clubs or crowds. § Adjusted for age-group, ethnicity, sexual orientation, educational qualifications, employment, and financial situation. \*\* uOR=Unadjusted odds ratio. aOR=adjusted odds ratio.

Sexual behaviours associated with HAV vaccination uptake included self-reported usage of chemsex-related recreational drugs in the previous year (aOR:1.87, (CI:1.16-3.02)), reporting ten or more male physical sex partners (aOR:1.85, (CI:1.4-2.44)), reporting attendance at SOP venues (aOR:1.58, (CI:1.22-2.05)), self-reporting ≥10 condomless anal sex (CAS) partners (aOR:1.44, (CI:1.12-1.86)) (all since Aug 2022). Health related factors associated with HAV vaccination uptake included self-reporting being a person living with HIV (PLWHIV) (aOR:2, (CI:1.38-2.88)), receiving a positive STI test in the past year (aOR:1.95, (CI:1.29-2.96)), visiting a SHS in the past year, (aOR:4.31, (CI:3.36-5.54)), STI test in the past year (aOR:3.25, (CI:2.53-4.18)) and reporting usage of HIV-PrEP since December 2021 (aOR:3.15, (CI:2.43-4.07)).

#### Hepatitis B Vaccination uptake

There was a bivariate association between HBV uptake and sexual orientation, educational qualifications, and employment status, see Table 3. There were lower odds of HBV vaccination uptake in those self-reporting as bisexual (aOR:0.57, (CI:0.39-0.82) vs gay/homosexual), having an education that was below degree-level (aOR:0.59, (CI:0.46-0.76)) and being unemployed (aOR:0.71, (CI:0.53-0.96)). There was no evidence of an independent association existing between HBV vaccination uptake and age however uptake was lower in both the 16-29 age group (aOR:0.44, (CI:0.3-0.65)) and the over 45 age group (aOR:0.53, (CI:0.4-0.7)).

**Table 3.** Sociodemographic characteristics and Health-related and sexual behaviour factors associated with self-reported HBV vaccination uptake among RiiSH-Mpox participants.

|  | n<br>total | % of RiSH-Mpox<br>participants | n of HBV<br>vaccinated<br>participants * | % of<br>group<br>vaccinated | % of HBV<br>vaccinate<br>d people | uOR ** | 95%<br>confidence<br>interval | P | aOR § | 95%<br>confidence<br>interval | P |
| --- | --- | --- | --- | --- | --- | --- | --- | --- | --- | --- | --- |
| **Sociodemographic Characteristics** |  |  |  |  |  |  |  |  |  |  |  |
| **Age-group** |  |  |  |  |  |  |  |  |  |  |  |
| 16-29 | 176 | 13 | 110 | 63 | 12 | 0.40 | 0.27 - 0.58 |  | 0.44 | 0.30 - 0.65 | 0.343 |
| 30-44 | 486 | 36 | 392 | 81 | 41 | 1.00 |  |  | 1.00 |  |  |
| 45+ | 671 | 50 | 452 | 67 | 47 | 0.49 | 0.38 - 0.65 | 0.343 | 0.53 | 0.40 - 0.70 |  |
| **Sexual orientation** |  |  |  |  |  |  |  |  |  |  |  |
| Gay/ homosexual | 1187 | 89 | 866 | 73 | 91 | 1.00 |  |  | 1.00 |  |  |
| Bisexual † | 146 | 11 | 88 | 60 | 9 | 0.56 | 0.39 - 0.80 | 0.002 | 0.57 | 0.39 - 0.82 | 0.002 |
| **Ethnicity** |  |  |  |  |  |  |  |  |  |  |  |
| White | 1223 | 92 | 872 | 71 | 91 | 1.00 |  |  | 1.00 |  |  |
| Other | 110 | 8 | 82 | 75 | 9 | 1.12 | 0.75 - 1.84 | 0.720 | 1.17 | 0.74 - 1.86 | 0.501 |
| **Country of Birth** |  |  |  |  |  |  |  |  |  |  |  |
| UK | 1113 | 83 | 796 | 72 | 83 | 1.00 |  |  | 1.00 |  |  |
| Other | 220 | 17 | 158 | 72 | 17 | 1.01 | 0.74 - 1.40 | 0.928 | 0.88 | 0.63 - 1.24 | 0.469 |
| **Country of Residence** |  |  |  |  |  |  |  |  |  |  |  |
| England | 1148 | 86 | 824 | 72 | 86 | 1.00 |  |  | 1.00 |  |  |
| Rest of UK | 185 | 14 | 130 | 70 | 14 | 1.01 | 0.74 - 1.40 | 0.928 | 0.95 | 0.67 - 1.35 | 0.770 |
| **Educational level** |  |  |  |  |  |  |  |  |  |  |  |
| Degree level or higher | 495 | 37 | 638 | 76 | 67 | 1.00 |  |  | 1.00 |  |  |
| Below degree level | 838 | 63 | 316 | 64 | 33 | 0.55 | 0.43 - 0.71 | <0.001 | 0.59 | 0.46 - 0.76 | <0.001 |
| **Employment** |  |  |  |  |  |  |  |  |  |  |  |
| Currently Employed | 1074 | 19 | 790 | 74 | 83 | 1.00 |  |  | 1.00 |  |  |
| Not Employed | 259 | 81 | 164 | 63 | 17 | 0.55 | 0.43 - 0.71 | <0.001 | 0.71 | 0.53 - 0.96 | 0.026 |
| **Household Composition** |  |  |  |  |  |  |  |  |  |  |  |
| Alone | 506 | 38 | 370 | 73 | 39 | 1.00 |  |  | 1.00 |  |  |
| With Others | 827 | 62 | 584 | 71 | 61 | 0.88 | 0.69 - 1.13 | 0.325 | 0.85 | 0.66 - 1.09 | 0.195 |
| **Relationship Status** |  |  |  |  |  |  |  |  |  |  |  |
| Relationship | 591 | 44 | 539 | 91 | 56 | 1.00 |  |  | 1.00 |  |  |
| Single | 742 | 56 | 415 | 56 | 44 | 1.13 | 0.89 - 1.43 | 0.330 | 1.12 | 0.88 - 1.44 | 0.360 |
| **Comfortable Financial Status** |  |  |  |  |  |  |  |  |  |  |  |
| Yes †† | 693 | 52 | 482 | 70 | 51 | 1.00 |  |  | 1.00 |  |  |
| No | 640 | 48 | 472 | 74 | 49 | 0.70 | 0.55 - 0.89 | 0.004 | 0.82 | 0.64 - 1.06 | 0.122 |
| **Health-related factors** |  |  |  |  |  |  |  |  |  |  |  |
| **HIV Status** |  |  |  |  |  |  |  |  |  |  |  |
| Negative/ Unknown | 1131 | 85 | 793 | 70 | 83 | 1.00 |  |  | 1.00 |  |  |
| PLWHIV | 202 | 15 | 161 | 80 | 17 | 1.67 | 1.16 - 2.41 | 0.006 | 1.84 | 1.26 - 2.68 | 0.001 |
| **MPOX-related behaviour modification ¶** |  |  |  |  |  |  |  |  |  |  |  |
| No | 626 | 47 | 434 | 69 | 45 | 1.00 |  |  | 1.00 |  |  |
| Yes | 707 | 53 | 520 | 74 | 55 | 1.23 | 0.97 - 1.56 | 0.088 | 1.14 | 0.90 - 1.46 | 0.281 |
| **STI Positive Test** |  |  |  |  |  |  |  |  |  |  |  |
| No | 1177 | 70 | 816 | 69 | 86 | 1.00 |  |  | 1.00 |  |  |
| Yes | 156 | 30 | 138 | 88 | 14 | 3.39 | 2.04 - 5.63 | <0.001 | 3.17 | 1.90 - 5.29 | <0.001 |
| **Visited SHC in past year** |  |  |  |  |  |  |  |  |  |  |  |
| No | 478 | 36 | 232 | 49 | 24 | 1.00 |  |  | 1.00 |  |  |
| Yes | 855 | 64 | 722 | 84 | 76 | 5.76 | 4.45 - 7.45 | <0.001 | 5.33 | 4.10 - 6.93 | <0.001 |
| **STI test in past year** |  |  |  |  |  |  |  |  |  |  |  |
| No | 405 | 30 | 198 | 49 | 21 | 1.00 |  |  | 1.00 |  |  |
| Yes | 928 | 70 | 756 | 81 | 79 | 4.60 | 3.56 - 5.93 | <0.001 | 4.29 | 3.31 - 5.58 | <0.001 |
| **PrEP Use in Past Year** |  |  |  |  |  |  |  |  |  |  |  |
| No | 734 | 55 | 428 | 58 | 45 | 1.00 |  |  | 1.00 |  |  |
| Yes | 599 | 45 | 526 | 88 | 55 | 5.15 | 3.87 - 0.69 | <0.001 | 4.68 | 3.51 - 6.26 | <0.001 |
| **Sexual behaviours** |  |  |  |  |  |  |  |  |  |  |  |
| **Usage of chemsex-related drugs in the previous year** |  |  |  |  |  |  |  |  |  |  |  |
| No | 1222 | 92 | 864 | 71 | 91 | 1.00 |  |  | 1.00 |  |  |
| Yes | 111 | 8 | 90 | 81 | 9 | 1.78 | 1.09 - 2.90 | 0.022 | 1.75 | 1.06 - 2.88 | 0.028 |
| **Number of Male Physical Sex Partners** |  |  |  |  |  |  |  |  |  |  |  |
| none | 100 | 8 | 64 | 64 | 7 | 1.00 |  |  | 1.00 |  |  |
| 1 | 180 | 14 | 103 | 57 | 11 | 0.75 | 0.45 - 1.25 |  | 0.71 | 0.43 - 1.19 | <0.001 |
| 2 to 4 | 389 | 29 | 268 | 69 | 28 | 1.25 | 0.79 - 1.98 |  | 1.16 | 0.72 - 1.86 |  |
| 5 to 9 | 280 | 21 | 208 | 74 | 22 | 1.63 | 1.00 - 2.65 |  | 2.51 | 0.92 - 2.48 |  |
| 10+ | 384 | 29 | 311 | 81 | 33 | 2.40 | 1.48 - 3.88 | <0.001 | 2.10 | 1.28 - 3.44 |  |
| **reported use of SOP venues in the lookback period** |  |  |  |  |  |  |  |  |  |  |  |
| No | 888 | 67 | 606 | 68 | 64 | 1.00 |  |  | 1.00 |  |  |
| Yes | 445 | 33 | 348 | 78 | 36 | 1.67 | 1.28 - 2.18 | <0.001 | 1.51 | 1.15 - 1.99 | 0.003 |
| **Number of CAS partners in the lookback period (since Dec 2021)** |  |  |  |  |  |  |  |  |  |  |  |
| none | 401 | 30 | 82 | 20 | 9 | 1.00 |  |  | 1.00 |  |  |
| 1 | 263 | 20 | 177 | 67 | 19 | 1.23 | 0.89 - 1.71 |  | 1.19 | 0.85 - 1.67 | 0.000 |
| 2 to 4 | 315 | 24 | 237 | 75 | 25 | 1.82 | 1.31 - 2.52 |  | 1.83 | 1.31 - 2.56 |  |
| 5 to 9 | 146 | 11 | 115 | 79 | 12 | 2.22 | 1.42 - 3.46 |  | 2.02 | 1.28 - 3.18 |  |
| 10+ | 208 | 16 | 174 | 84 | 18 | 3.06 | 2.01 - 4.65 | 0.000 | 2.91 | 1.89 - 4.47 |  |

\* Participants with self-reported uptake of $\geq 1$ HBV vaccine dose(s). † Includes those identifying as bisexual, straight, or another way. †† Top two quartiles ("I am comfortable"/"I am very comfortable" from the question, "How would you best describe your current financial situation". ¶ Behaviour modification includes self-report of any of the following from May 2022: fewer sexual partners, reduced visits to sex on premises venues or PSE (i.e. cruising grounds), and avoiding: all sex, condomless anal sex, skin-to-skin contact, or visiting clubs or crowds. § Adjusted for age-group, ethnicity, sexual orientation, educational qualifications, employment, and financial situation. \*\* uOR=Unadjusted odds ratio. aOR=adjusted odds ratio.

Sexual behaviours associated with uptake included reported usage of chemsex-related recreational drugs in the previous year (aOR:1.75, (CI:1.06-2.88)), reporting ten or more physical male sex partners (aOR:1.89, (CI:1.41-2.53)) reporting attendance at SOP venues (aOR:1.51, (CI:1.15-1.99)), self-reporting ≥10 CAS partners (aOR:1.77, (CI:1.37-2.29)) (all since Aug 2022). There were associations with reporting being a PLWHIV (aOR:1.84, (CI:1.26-2.68)), receiving a positive STI test in the past year (aOR:3.17, (CI:1.9-5.29)), visiting a SHS in the past year, (aOR:5.33, (CI:4.1-6.93)), having an STI test in the past year (aOR:4.29, (CI:3.31-5.58)) and reporting usage of HIV-PrEP since Dec 2021 (aOR:4.68, (CI:3.51-6.26)).

#### HPV vaccination uptake

Among eligible individuals the only sociodemographic factors found to have an association with HPV vaccination uptake were being in the 16-29 (younger) age-category (aOR:0.62, (CI:0.43-0.89 vs those aged 30-44)) and having an education below degree-level (aOR:0.55, (CI:0.39-0.78)), see Table 4.

**Table 4.**
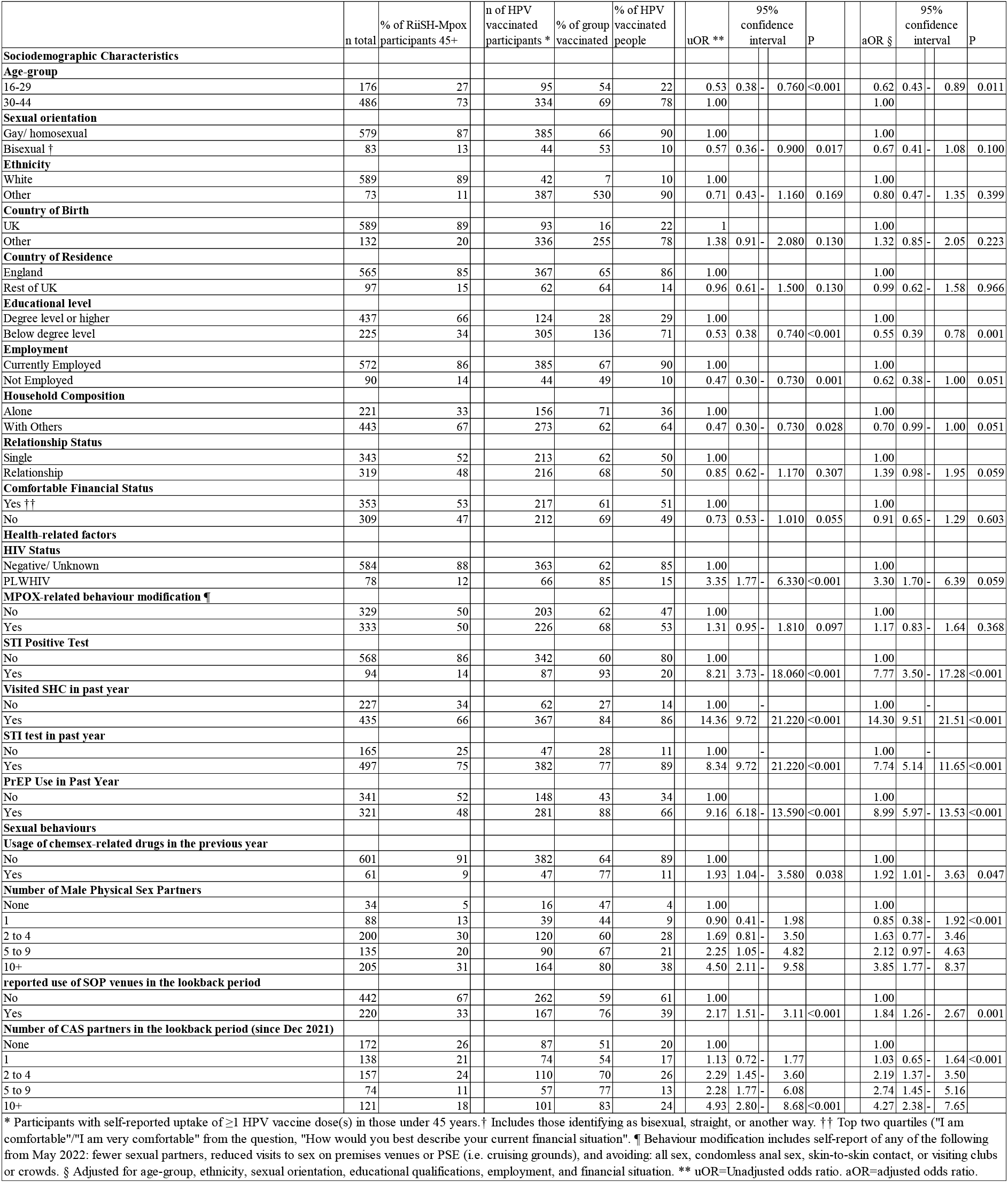
Sociodemographic characteristics and Health-related and sexual behaviour factors associated with self-reported HPV vaccination uptake among RiiSH-Mpox participants below 45.

The sexual behaviours that were associated with HPV vaccination uptake include usage of chemsex-related drugs in the previous year (aOR:1.92, (CI:1.01-3.63)), reporting ten or more physical male sex partners (aOR:3.85, (CI:1.77-8.37)) reporting attendance at SOP venues (aOR:1.84, (CI:1.26-2.67)), self-reporting ≥10 CAS partners (aOR:4.27, (CI:2.38-7.65)) (all since August 2022). There were increased odds of HPV vaccination in a multivariable model amongst those reporting being a PLWHIV (aOR:3.3, (CI:1.7-6.39)), receiving a positive STI test in the past year (aOR:7.77, (CI:3.5-17.28)), visiting a SHS in the past year (aOR:14.3, (CI:9.51-21.51)), STI test in the past year (aOR:7.74, (CI:5.14-11.65)) and reporting usage of HIV-PrEP since December 2021 (aOR:4.12, (aOR:8.99, (CI:5.97-13.53)).

## Discussion

Analyses of a large, community-based survey of GBMSM in the UK show that vaccination uptake was high but far from universal for all targeted vaccinations, with uptake at around two-thirds across each of the STI/VH vaccinations considered. In GBMSM attending SHS, approximately a quarter were considered potential missed opportunities for vaccination across each vaccine type. There was highly correlated uptake between different vaccines and similar sociodemographic, behavioural and SHS use factors were associated with uptake of each STI/VH vaccine. This provides an important insight into HAV, HBV and HPV vaccination uptake amongst GBMSM for which there is limited uptake data in the UK^21^.

The sociodemographic factors identified in this study found to commonly be associated with lower vaccination uptake were younger age, self-identifying as bisexual, and markers of lower financial or social capital such as an education below degree-level or being unemployed. Vaccination uptake was higher amongst those reporting sexual behaviour which increases risk of STI transmission, and relatedly, in those reporting recent SHS use (greater sexual behaviour is often associated with SHS utilisation^20^); indicating that those who are most at need of vaccines are receiving them. This mirrors findings of a previous analysis of mpox vaccination uptake^15^. Vaccine completion ranged from 42% for mpox to 89% for HAV. The lower completion rate for the mpox vaccination is likely due to the recency of implementation of the vaccination in response to the outbreak in 2022, as well as the fact that the offer of second doses only commenced 2-3 months prior to the survey^10^.

The lower odds of vaccination amongst younger age groups has previously been reported in other studies assessing HBV and HPV vaccination^16,19,21^. Gay self-identification, higher levels of education and employment were also consistently found to be linked with higher vaccination uptake amongst GBMSM, including both HBV and HPV vaccinations^16,18,21^. Young people and bisexual/straight-identifying GBMSM have been identified as groups that have a higher sexual risk and a lower SHS engagement^22–24^. Promotion focused on bisexual men is important to reduce the burden of these vaccine preventable STI/VHs amongst GBMSM, but also potentially amongst women^25^.

There is also a consistent association between higher vaccination uptake and SHS usage (although HAV/HBV vaccines are also administered outside of SHS). Previous research has found that GBMSM are not actively seeking out vaccines such as the HPV vaccine^11,17^ and that vaccination for HPV is reliant on recommendation by clinicians^26,27^. It follows that to become aware of, and to be identified as a candidate for vaccination, they must be attending SHSs, (HPV vaccination is targeted to GBMSM attending SHS only). This association has been identified previously with one study finding that 90.9% of GBMSM reporting HPV vaccination had attended a SHS in the previous year^28^ (86% in this sample).

Those with markers of lower financial or social capital reported lower vaccination uptake, indicating that health inequalities faced by GBMSM are exacerbated amongst those who already experience inequity. The effects of marginalisation may be compounded, and this requires an intersectional approach to understanding these groups and how to target them effectively. Understanding their behaviour and whether they require changes in policy and tailored interventions to increase their engagement with SHS, and subsequently uptake of STI/VH vaccinations, is needed. This could be through health-promotion messaging such as community outreach programmes and media-based advertisements to reach those who may not normally access SHS. One means may be through encouraging online SHS^29^ to identify potential vaccination candidates, but this does require online services that are well-integrated with physical SHS and have good signposting for GBMSM, as vaccination requires attendance at a physical venue.

Vaccine completion is also incredibly important to confer the maximum protection against infection, amongst this sample many of those that initiated vaccination completed the course of vaccination. One previous study assessing incomplete vaccination amongst GBMSM in England found that the most common reason for not receiving additional vaccine doses was because they were unaware when their next dose was due^26^. This indicates that vaccine completion requires additional or alternative promotion to encourage that those who receive their initial dose will continue to receive additional vaccine doses.

This study has limitations due to the cross-sectional methodology used. For example, there may have been recall error as vaccination was self-reported. Also, participants’ behaviours may have changed, and chronology of vaccination relative to behaviour cannot be ascertained (except vaccination for mpox which only became established June 2022^10^). There may also be a degree of misclassification bias by retrospectively applying eligibility criteria to participants as their behaviours may have changed over time.

Due to the questions regarding vaccination which fell into the ‘ever’ lookback period, it is impossible to know when vaccination occurred, which could explain high association between HAV and HBV vaccination due to combined packaging of the HAV and HBV vaccination or administration of the combined (HAV/HBV) Twinrix vaccine.

There are also limitations based on the participants’ demographics with lower response numbers from young people and people who did not self-describe as white. This necessitated dichotomising variables, limiting the quality of these findings for minority groups who also have an increased burden of STIs^2,30,31^, using dichotomised measures limits this study’s ability to assess inequalities in vaccine access by ethnicity or gender identity. There is also potential participation bias, for example the sample appears to be more health conscious than the general GBMSM population. For instance, HPV vaccine uptake within the eligible sample was 65% compared to 34% of GBMSM nationally^12^. This sample also excludes GBMSM not using the applications that this survey was advertised on. However, unlike previous studies^28,32^ which often focus on those attending SHS, the target population for this study was the GBMSM community more broadly.

Improving uptake is dependent on understanding barriers faced by GBMSM which limit vaccination uptake. Within the UK, awareness of factors affecting uptake (especially as the JCVI has advised routine implementation of opportunistic vaccination for mpox^33^ and gonorrhoea [the latter using 4CMenB vaccination^34^]), will help inform practice and health promotion that aligns with NHS^35^ and UKHSA^36^ health equity goals. In an international context, these results could raise awareness of potential limitations in vaccination uptake due to service provision differences and sociodemographic factors in equivalent and future GBMSM-selective vaccination programmes.

## Data Availability

The data that support the findings of this study are available upon reasonable request from the UK Health Security Agency (UKHSA). Requests can be directed to Dr Hamish Mohammed.

## Acknowledgements

The authors wish to thank all the participants who took part in this study. We acknowledge members of the National Institute for Health and Care Research Health Protection Research Unit (NIHR HPRU) in Blood Borne and Sexually Transmitted Infections (BBSTI) Steering Committee: Professor Caroline Sabin (HPRU Director), Dr John Saunders (UK Health Security Agency Lead), Professor Catherine Mercer, Dr Hamish Mohammed (previously Professor Gwenda Hughes), Professor Greta Rait, Dr Ruth Simmons, Professor William Rosenberg, Dr Tamyo Mbisa, Professor Rosalind Raine, Dr Sema Mandal, Dr Rosamund Yu, Dr Samreen Ijaz, Dr Fabiana Lorencatto, Dr Rachel Hunter, Dr Kirsty Foster and Dr Mamooma Tahir. The authors would also like to thank Takudzwa Mukiwa (Terrence Higgins Trust) and Will Nutland (The Love Tank) for their review of the mpox module in the RiiSH-Mpox survey used for this research.

---

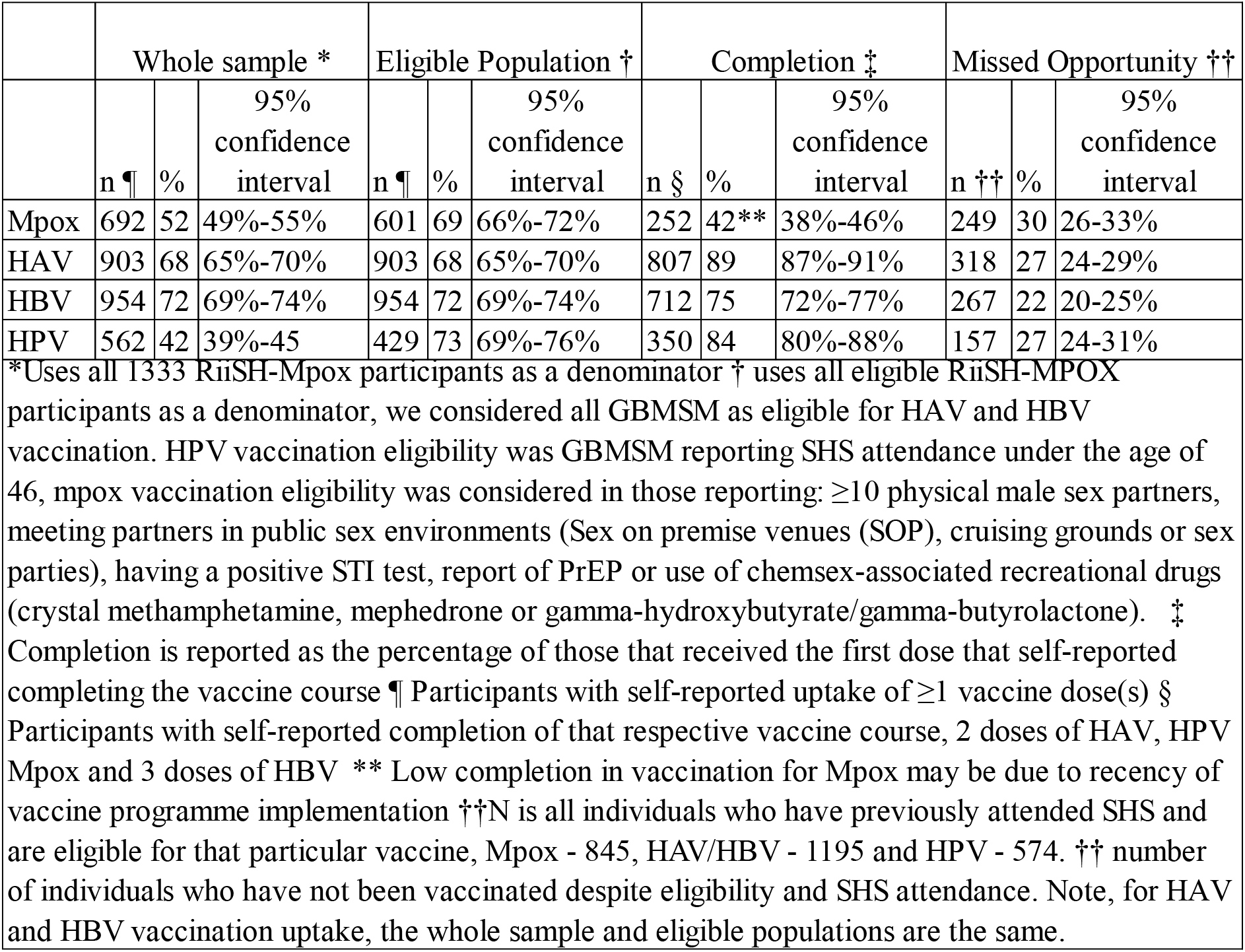

|  | Whole sample * |  |  | Eligible Population † |  |  | Completion ‡ |  |  | Missed Opportunity †† |  |  |
| --- | --- | --- | --- | --- | --- | --- | --- | --- | --- | --- | --- | --- |
|  | n ¶ | % | 95% confidence interval | n ¶ | % | 95% confidence interval | n § | % | 95% confidence interval | n †† | % | 95% confidence interval |
| Mpox | 692 | 52 | 49%-55% | 601 | 69 | 66%-72% | 252 | 42** | 38%-46% | 249 | 30 | 26-33% |
| HAV | 903 | 68 | 65%-70% | 903 | 68 | 65%-70% | 807 | 89 | 87%-91% | 318 | 27 | 24-29% |
| HBV | 954 | 72 | 69%-74% | 954 | 72 | 69%-74% | 712 | 75 | 72%-77% | 267 | 22 | 20-25% |
| HPV | 562 | 42 | 39%-45 | 429 | 73 | 69%-76% | 350 | 84 | 80%-88% | 157 | 27 | 24-31% |

\*Uses all 1333 RiSH-Mpox participants as a denominator † uses all eligible RiSH-MPOX participants as a denominator, we considered all GBMSM as eligible for HAV and HBV vaccination. HPV vaccination eligibility was GBMSM reporting SHS attendance under the age of 46, mpox vaccination eligibility was considered in those reporting: ≥10 physical male sex partners, meeting partners in public sex environments (Sex on premise venues (SOP), cruising grounds or sex parties), having a positive STI test, report of PrEP or use of chemsex-associated recreational drugs (crystal methamphetamine, mephedrone or gamma-hydroxybutyrate/gamma-butyrolactone). ‡ Completion is reported as the percentage of those that received the first dose that self-reported completing the vaccine course ¶ Participants with self-reported uptake of ≥1 vaccine dose(s) § Participants with self-reported completion of that respective vaccine course, 2 doses of HAV, HPV Mpox and 3 doses of HBV \*\* Low completion in vaccination for Mpox may be due to recency of vaccine programme implementation ††N is all individuals who have previously attended SHS and are eligible for that particular vaccine, Mpox - 845, HAV/HBV - 1195 and HPV - 574. †† number of individuals who have not been vaccinated despite eligibility and SHS attendance. Note, for HAV and HBV vaccination uptake, the whole sample and eligible populations are the same.

**Appendix A.** Sociodemographic, Health-related and sexual behaviour characteristics of those who self-reported ≥1 dose of the Mpox, HAV, HBV and HPV vaccines amongst RiiSH-Mpox participants.

| Appendix A. Sociodemographic, Health-related and sexual behaviour characteristics of those who self-reported ≥1 dose of the Mpox, HAV, HBV and HPV vaccines amongst RiSH-Mpox participant |  |  |  |  |  |  |  |  |  |  |
| --- | --- | --- | --- | --- | --- | --- | --- | --- | --- | --- |
|  | n total | % of RiSH-Mpox participants | n of mpox vaccinated participants | % of MPOX vaccinated people | n of HAV vaccinated participants | % of HAV vaccinated people | n of HBV vaccinated participants | % of HBV vaccinated people | n of HPV vaccinated participants | % of HPV vaccinated people |
| **Sociodemographic Characteristics** |  |  |  |  |  |  |  |  |  |  |
| **Age-group** |  |  |  |  |  |  |  |  |  |  |
| 16-29 | 176 | 13 | 78 | 11 | 101 | 11 | 110 | 12 | 95 | 17 |
| 30-44 | 486 | 36 | 263 | 38 | 358 | 40 | 392 | 41 | 334 | 59 |
| 45+ | 671 | 50 | 351 | 51 | 444 | 49 | 452 | 47 | 133 | 24 |
| **Sexual orientation** |  |  |  |  |  |  |  |  |  |  |
| Gay/ homosexual | 1187 | 89 | 645 | 93 | 823 | 91 | 866 | 91 | 509 | 91 |
| Bisexual * | 146 | 11 | 47 | 7 | 80 | 9 | 88 | 9 | 53 | 9 |
| **Ethnicity** |  |  |  |  |  |  |  |  |  |  |
| White | 1223 | 92 | 635 | 92 | 832 | 92 | 872 | 91 | 512 | 91 |
| Other | 110 | 8 | 57 | 8 | 71 | 8 | 82 | 9 | 50 | 9 |
| **Country of Birth** |  |  |  |  |  |  |  |  |  |  |
| UK | 1113 | 83 | 566 | 82 | 756 | 84 | 796 | 83 | 446 | 79 |
| Other | 220 | 17 | 126 | 18 | 147 | 16 | 158 | 17 | 116 | 21 |
| **Country of Residence** |  |  |  |  |  |  |  |  |  |  |
| England | 1148 | 86 | 613 | 89 | 779 | 86 | 824 | 86 | 479 | 85 |
| Rest of UK | 185 | 14 | 79 | 11 | 124 | 14 | 130 | 14 | 83 | 15 |
| **Educational level** |  |  |  |  |  |  |  |  |  |  |
| Degree level or higher | 838 | 63 | 199 | 29 | 594 | 66 | 638 | 67 | 394 | 70 |
| Below degree level | 495 | 37 | 493 | 71 | 309 | 34 | 316 | 33 | 169 | 30 |
| **Employment** |  |  |  |  |  |  |  |  |  |  |
| Currently Employed | 1074 | 19 | 595 | 86 | 749 | 83 | 790 | 83 | 497 | 88 |
| Not employed | 259 | 81 | 97 | 14 | 154 | 17 | 164 | 17 | 65 | 12 |
| **Household Composition** |  |  |  |  |  |  |  |  |  |  |
| With others | 827 | 38 | 515 | 74 | 351 | 39 | 584 | 61 | 342 | 61 |
| Alone | 506 | 62 | 278 | 40 | 552 | 61 | 370 | 39 | 220 | 39 |
| **Relationship Status** |  |  |  |  |  |  |  |  |  |  |
| In relationship | 591 | 44 | 320 | 46 | 510 | 56 | 539 | 56 | 291 | 52 |
| Single | 742 | 56 | 372 | 54 | 393 | 44 | 415 | 44 | 271 | 48 |
| **Comfortable Financial Status** |  |  |  |  |  |  |  |  |  |  |
| Yes | 693 | 52 | 331 | 48 | 453 | 50 | 482 | 51 | 282 | 50 |
| No | 640 | 48 | 361 | 52 | 450 | 50 | 472 | 49 | 280 | 50 |
| **Sexual behaviours** |  |  |  |  |  |  |  |  |  |  |
| **Usage of chemsex-related drugs in the previous year** |  |  |  |  |  |  |  |  |  |  |
| No | 1222 | 92 | 617 | 89 | 815 | 90 | 864 | 91 | 500 | 89 |
| Yes | 111 | 8 | 75 | 11 | 88 | 10 | 90 | 9 | 62 | 11 |
| **Number of Male Physical Sex Partners** |  |  |  |  |  |  |  |  |  |  |
| None | 100 | 8 | 31 | 4 | 64 | 7 | 64 | 7 | 28 | 5 |
| 1 | 180 | 14 | 41 | 6 | 93 | 10 | 103 | 11 | 47 | 8 |
| 2 to 4 | 389 | 29 | 170 | 25 | 256 | 28 | 268 | 28 | 159 | 28 |
| 5 to 9 | 280 | 21 | 180 | 26 | 192 | 21 | 208 | 22 | 119 | 21 |
| 10+ | 384 | 29 | 270 | 39 | 298 | 33 | 311 | 33 | 209 | 37 |
| **reported use of SOP venues in the lookback period** |  |  |  |  |  |  |  |  |  |  |
| No | 888 | 67 | 382 | 55 | 568 | 63 | 606 | 64 | 346 | 62 |
| Yes | 445 | 33 | 310 | 45 | 335 | 37 | 348 | 36 | 216 | 38 |
| **Number of CAS partners in the lookback period** |  |  |  |  |  |  |  |  |  |  |
| None | 401 | 30 | 57 | 8 | 245 | 27 | 82 | 9 | 119 | 21 |
| 1 | 263 | 20 | 99 | 14 | 159 | 18 | 177 | 19 | 96 | 17 |
| 2 to 4 | 315 | 24 | 185 | 27 | 219 | 24 | 237 | 25 | 143 | 25 |
| 5 to 9 | 146 | 11 | 109 | 16 | 118 | 13 | 115 | 12 | 78 | 14 |
| 10+ | 208 | 16 | 143 | 21 | 162 | 18 | 174 | 18 | 126 | 22 |
| **Health-related factors** |  |  |  |  |  |  |  |  |  |  |
| **HIV Status** |  |  |  |  |  |  |  |  |  |  |
| Negative/ Unknown | 1131 | 85 | 575 | 83 | 744 | 82 | 793 | 83 | 461 | 82 |
| PLWHIV | 202 | 15 | 117 | 17 | 159 | 18 | 161 | 17 | 101 | 18 |
| **MPOX-related behaviour modification ††** |  |  |  |  |  |  |  |  |  |  |
| No | 626 | 47 | 247 | 36 | 412 | 46 | 434 | 45 | 264 | 47 |
| Yes | 707 | 53 | 445 | 64 | 491 | 54 | 520 | 55 | 298 | 53 |
| **STI Positive Test** |  |  |  |  |  |  |  |  |  |  |
| No | 1177 | 70 | 568 | 82 | 778 | 86 | 816 | 86 | 457 | 81 |
| Yes | 156 | 30 | 124 | 18 | 125 | 14 | 138 | 14 | 105 | 19 |
| **Visited SHC in past year** |  |  |  |  |  |  |  |  |  |  |
| No | 478 | 36 | 82 | 12 | 221 | 24 | 232 | 24 | 60 | 11 |
| Yes | 855 | 64 | 610 | 88 | 682 | 76 | 722 | 76 | 484 | 86 |
| **STI test in past year** |  |  |  |  |  |  |  |  |  |  |
| No | 405 | 30 | 81 | 12 | 196 | 22 | 198 | 21 | 60 | 11 |
| Yes | 928 | 70 | 611 | 88 | 707 | 78 | 756 | 79 | 502 | 89 |
| **PrEP Use in Past Year** |  |  |  |  |  |  |  |  |  |  |
| No | 734 | 55 | 228 | 33 | 415 | 46 | 428 | 45 | 201 | 36 |
| Yes | 599 | 45 | 464 | 67 | 488 | 54 | 526 | 55 | 361 | 64 |

\* also includes those identifying straight, or another way. . † Top two quartiles ("I am comfortable"/"I am very comfortable" from the question, "How would you best describe your current financial situation". †† Behaviour modification includes self-report of any of the following: fewer sexual partners, reduced visits to sex on premises venues or PSE and avoiding: all sex, condomless anal sex, skin-to-skin contact, or visiting clubs or crowds from May 2022.

**Appendix B.** Vaccine uptake and association between uptake of different STI vaccine types amongst RiiSH-Mpox participants.

|  |  |  |  |  |  |  |  |  |  |  |  |  |  |
| --- | --- | --- | --- | --- | --- | --- | --- | --- | --- | --- | --- | --- | --- |
|  | n | n receiving both vaccines * | % receiving both vaccines | uOR † | 95% confidence interval |  |  | P | aOR † † | 95% confidence interval |  |  | P |
| Any Vaccine Uptake | 1135 |  | 97 |  |  |  |  |  |  |  |  |  |  |
| Mpox Vaccine uptake |  |  |  |  |  |  |  |  |  |  |  |  |  |
| Mpox | 692 |  |  |  |  |  |  |  |  |  |  |  |  |
| HAV | 903 | 558 | 62 | 3.57 | 2.80 | - | 4.56 | <0.001 | 3.30 | 2.57 | - | 4.25 | <0.001 |
| HBV | 954 | 581 | 61 | 3.76 | 2.91 | - | 4.86 | <0.001 | 3.40 | 2.61 | - | 4.42 | <0.001 |
| HPV | 562 | 387 | 69 | 3.38 | 2.69 | - | 4.25 | <0.001 | 4.14 | 3.16 | - | 5.42 | <0.001 |
| HAV Vaccine Uptake | 903 |  | 68 |  |  |  |  |  |  |  |  |  |  |
| Mpox | 692 | 558 | 81 | 3.57 | 2.80 | - | 4.56 | <0.001 | 3.30 | 2.57 | - | 4.25 | <0.001 |
| HAV | 903 |  |  |  |  |  |  |  |  |  |  |  |  |
| HBV | 954 | 794 | 83 | 12.29 | 9.29 | - | 16.26 | <0.001 | 11.98 | 9.01 | - | 15.92 | <0.001 |
| HPV | 562 | 490 | 87 | 5.90 | 4.44 | - | 7.84 | <0.001 | 8.04 | 5.82 | - | 11.11 | <0.001 |
| HBV Vaccine uptake | 954 |  | 72 |  |  |  |  |  |  |  |  |  |  |
| Mpox | 692 | 581 | 84 | 3.76 | 2.91 | - | 4.86 | <0.001 | 3.40 | 2.61 | - | 4.42 | <0.001 |
| HAV | 903 | 794 | 88 | 12.29 | 9.29 | - | 16.26 | <0.001 | 11.98 | 9.01 | - | 15.92 | <0.001 |
| HBV | 954 |  |  |  |  |  |  |  |  |  |  |  |  |
| HPV | 562 | 512 | 91 | 7.62 | 5.15 | - | 10.53 | <0.001 | 9.27 | 6.51 | - | 13.21 | <0.001 |
| HPV Vaccine Uptake |  |  | 42 |  |  |  |  |  |  |  |  |  |  |
| Mpox | 692 | 387 | 56 | 3.38 | 2.69 | - | 4.25 | <0.001 | 3.30 | 2.57 | - | 4.25 | <0.001 |
| HAV | 903 | 490 | 54 | 5.90 | 4.44 | - | 7.84 | <0.001 | 8.04 | 5.82 | - | 11.11 | <0.001 |
| HBV | 954 | 512 | 54 | 7.62 | 5.52 | - | 10.53 | <0.001 | 9.27 | 6.51 | - | 13.21 | <0.001 |
| HPV | 562 |  |  |  |  |  |  |  |  |  |  |  |  |

\* Participants with self-reported uptake of ≥1 vaccine dose of both vaccine types.† uOR=Unadjusted odds ratio. aOR=adjusted odds ratio. † † Adjusted for age-group, sexual orientation, educational level, employment, ethnicity and financial situation.

